# Self-assessment and rest-activity rhythm monitoring for effective bipolar disorder management: a longitudinal actigraphy study

**DOI:** 10.1101/2025.03.11.25323782

**Authors:** Bianca Pfaffenseller, Jakub Schneider, Taiane de Azevedo Cardoso, Mario Simjanoski, Martin Alda, Flavio Kapczinski, Eduard Bakstein

## Abstract

**Background:** Recurrent course and disruption of circadian rhythms are among the core features of bipolar disorder (BD). Thus, ongoing symptom monitoring is an essential part of good clinical management.

**Objective:** We conducted a study to validate the English version of the ASERT (Aktibipo questionnaire), a tool for self-assessment of mood symptoms. We also analyzed the relationship of self-assessed symptoms with clinician ratings and actigraphy measures, and investigated the possibility of predicting depressive episodes using subjective and digital measures.

**Methods:** This was a longitudinal study of individuals with BD, followed for up to 11 months. The participants completed weekly mood self-assessments (ASERT) using a smartphone app and wore wrist actigraphs. During monthly appointments, the severity of their mood symptoms was rated by clinicians, and the participants completed questionnaires addressing overall functioning (FAST), and biological rhythms (BRIAN).

**Results:** The study confirmed the validity and reliability of the ASERT as a measure of subjective mood. Additionally, we found significant associations between ASERT responses, clinical scales, and actigraphy data. In our analysis, a combination of self-assessment and actigraphy data detected depression relapse with 67% sensitivity, 90% specificity, and 81% balanced accuracy. Furthermore, we observed a strong correlation between the stability of daily routine and overall functioning, emphasizing the significance of circadian rhythm disruptions in BD.

**Conclusion:** This study highlights the potential of digital tools, such as digitally administered self-assessments and actigraphy, to enhance the management of BD by providing valuable insights into mood states and detecting relapse. Further research is needed to refine and optimize these tools for widespread clinical application, such as informing personalized treatment plans.

## Introduction

Bipolar disorder (BD) affects approximately 1-2% of the world population, profoundly impacting the quality of life, functioning, and overall health^1^. Increasing evidence strongly suggests a pivotal role for circadian rhythms in the onset, course, and management of BD^2–4^. Sleep and circadian disruption could also be found in unaffected individuals at risk of BD^5^. as well as early in the course of the illness^6^. Even during remission, patients may experience dysregulation of circadian rhythm and sleep, including reduced overall activity and disrupted sleep patterns^7–10^. Importantly, dysregulation of circadian rhythms has been associated with illness recurrences^11^. Monitoring variables related to circadian rhythms, such as sleep and activity, can inform BD prognosis and contribute to more effective illness management^12^.

There is a growing interest in digital tools that can assist in the clinical management and self-management of psychiatric disorders^13,14^. Digitally administered self-assessment questionnaires (Ecological Momentary Assessments - EMA) are one example of such a tool^15^. Such short questionnaires are easy to complete, allowing for frequent data collection. EMA can be even more effective when used with a device such as an actigraph, which continuously monitors daily routines^16^. This approach enables patients to identify personal risk factors that may trigger symptom worsening^11,17^. By recognizing and avoiding or minimizing these risk factors, patients can actively manage their condition and seek timely help. Considering that sleep and activity parameters are reliable indicators of mood disruption associated with relapses and can be objectively assessed at a low cost, their implementation in clinical practice for monitoring the clinical course of BD is highly feasible.

The relapse rate in BD is relatively high; a recent large study showed that 25.5% of individuals with BD experienced at least one relapse within a 5-year period, with 39.1% of those experiencing multiple relapses^18^. And in a meta-analysis of naturalistic studies, the recurrence rate was up to 26.3% per year^19^.

In a previous study^20^, we introduced a novel questionnaire designed for remote mood monitoring, the ASERT (Aktibipo SElf-RaTing questionnaire). The ASERT rates subjective mood in patients with BD. Implemented as a smartphone application, it is a brief and user-friendly tool evaluating a variety of mood symptoms. In a previous study, we have demonstrated its effectiveness in detecting symptom changes over time^20^.

The current study had three objectives: (1) validate the English version of ASERT, (2) examine the relationship between ASERT, clinician symptoms ratings, and actigraphy variables, and (3) quantify the contribution of subjective and objective measures to the detection of BD recurrences. To address these objectives, we collected long-term actigraphy data using an actigraphy wristband and subjective self-reported mood data using a smartphone app. Additionally, clinician-rated scales were used to assess the severity of symptoms, overall functioning, and biological rhythms. This comprehensive approach allows to identify variables relevant to the clinical course and inform data-driven clinical decision-making in BD.

The present study addressed the following questions:

(1a) Does the ASERT questionnaire reliably measure BD mood symptoms compared to standard clinician-rated scales (Convergent Validity)?

(1b) Does the English version of the ASERT maintain its originally described internal structure (Structural Validity)?

(2) What is the relationship between ASERT self-report responses, clinician ratings, and actigraphy parameters?

(3) Can the ASERT questionnaire, alone or in combination with actigraphy data, detect clinical episodes (e.g., depression relapse)?

## Methods

### Study design, setting and participants

This cohort study consisted of an observational longitudinal approach with up to 11 months of follow-up to monitor a wide array of metrics, including sleep, physical activity, circadian rhythmicity, weekly mood self-records and monthly assessments of the severity of mood symptoms, clinical status, including hospitalizations, biological rhythms, and psychosocial functioning. A convenience sampling was adopted, and participants were recruited at the outpatient Mood Disorders Clinic at St. Joseph Healthcare Hamilton, a teaching psychiatric hospital affiliated with McMaster University, Hamilton, Canada, and through online advertisements on Facebook and Instagram profiles of the research group. The study was conducted remotely, and research interviews were conducted online using the Zoom platform. The inclusion criteria were: (1) diagnosis of BD by a psychiatrist, (2) remission at baseline confirmed by the Montgomery–Aasberg Depression Rating Scale^21^ (MADRS) and the Young Mania Rating Scale^22^ (YMRS) (MADRS total score ≤ 9 and YMRS total score ≤ 12), and (3) age between 18 and 70 years. Exclusion criteria included (1) cognitive impairment interfering with the ability to consent and follow the study protocol and (2) organic psychiatric disorder. Research personnel obtained informed consent from participants following an approved protocol by the local Research Ethics Board (Hamilton Integrated Research Ethics Board (HiREB), Project #13581). Participants were compensated with a total of $40.00 for their involvement in the study. The planned sample size (of 20 participants) is in line with many previous actigraphy studies^10^, most of which had shorter follow-up compared to the current study.

### Clinical assessments

At baseline, we collected sociodemographic and clinical characteristics of the participants, including sex, age, level of education, number of lifetime mood episodes, age at onset, current medications, number of psychiatric hospitalizations and history of suicide attempts, and we rated the severity of mood symptoms using the YMRS and the MADRS. The participants also completed the Functioning Assessment Short Test^23^ (FAST), a 24-item assessment including 6 domains of function (autonomy, occupational functioning, cognitive functioning, financial, interpersonal, and leisure time), and the Biological Rhythms Interview of Assessment in Neuropsychiatry^24^ (BRIAN), a 21-item assessment evaluating rhythm disturbance in sleep, activity, social and eating patterns. Following the first clinical interview, monthly online visits included YMRS and MADRS ratings, FAST and BRIAN. All assessments and symptom ratings were performed by properly trained research personnel at the Mood Disorders Clinic at St. Joseph Healthcare Hamilton, Canada.

### Continuous data collection

The collection of continuous data was performed using the Mindpax platform, consisting of a mobile application installed on the participants’ smartphones and a proprietary actigraphy wristband (Mindpax). Participants received the actigraphs at the beginning of the study and linked them to their accounts in the mobile application. The same application was used to collect the ASERT self-assessments on a weekly basis.

#### Subjective quantification of participant’s mood

During the follow-up, the participants completed weekly self-reports using the ASERT questionnaire^20^. The ASERT questionnaire includes four items indexing depression (items 1 to 4), four items focused on mania (items 5 to 8), and two non-specific items - inner tension (item 9), and inability to focus (item 10) over the previous week (Table S2). We used an English version of the questionnaire, validated using a back-translation of the original version^20^.

#### Objective assessment of the participant’s sleep and activity pattern

The participants were instructed to wear the actigraphy wristband on the wrist of the non-dominant arm and remove it only when necessary. The battery life is sufficiently long and does not require any charging during the study. The measured acceleration was aggregated over 30-second epochs and transmitted through the Mindpax.me mobile application to a secure database, where activity and sleep-related metrics were calculated.

Actigraphy parameters were calculated using proprietary scripts in Mathworks Matlab^25^. Cosinor analysis, obtained from 7- and 14-day windows, was used to estimate the mesor (the overall activity) associated with BD and depression^26–28^, amplitude (representing the sleep-wake differences) and acrophase (timing of daily activity and rest), associated with chronotype, both as state and trait marker^29^. Non-parametric analysis, based on 7- and 14-day windows, was used to calculate intradaily variability (IV) (corresponding to the fragmentation of the daily activity profile), which was found to be associated with sleep quality, cognitive and motor performance, as well as with social interactions^30^, and interdaily stability (IS) (corresponding to the stability of the daily activity profile across days), which has been used as a measure of synchronization to the light-dark cycle and cognitive functions^7,30^, using an actigraphy data resampled to 20-min averages, which has shown to provide the most stable results^31^. Sleep epochs were extracted using the Mindpax proprietary algorithm, and the daily sleep duration was calculated as the sum of durations of all detected sleep epochs within 24 hours. From the daily sleep duration, the window average, and the median absolute differences (MAD) were calculated in 7- and 14-day windows to measure the overall sleep duration and variability thereof.

### Statistical Analysis

After exploratory data analysis, the **convergent validity** of the depression and mania subscores of the ASERT was measured by comparison to the respective clinical scales (MADRS total scores, YMRS total scores). Each clinical scale was matched to the nearest ASERT response for a given participant within a 15-day (+/-7 days) time window. The dependency was tested using the linear mixed-effects models (LME), with the MADRS/YMRS scores as the dependent variable and the respective ASERT subscore as the fixed effect predictor variable, with random intercepts for individual participants.

Next, we compared ASERT subscores with the FAST and BRIAN total scores, measuring functional and circadian impairment, respectively. Finally, we compared the ASERT subscores with the BRIAN circadian rhythm subscore, which was evaluated separately and not included in the BRIAN total score, as it measures the predominant chronotype rather than the circadian disruption.

The choice of LME models over standard linear models was based on the longitudinal character of the study, in which subsequent measurements in the same participant can not be assumed independent. Therefore, the LME includes the fixed effect, quantifying the overall dependency, and random effects, quantifying deviations of the individual participants. In all comparisons, we used the models with random intercepts, as models combining random intercept and slope faced non-convergence issues and provided minimal to no improvement over the random intercept-only models. Data analysis and statistics were performed using the R statistical software^32^. The LME models were performed using the lme4 package^33^. The statistical inference on the LME fixed effects was performed using Satterthwaite’s method, which has been shown to provide accurate estimates^34^ and which is implemented in the lmerTest package^35^.

For evaluation of convergent validity and comparison with objective data and clinical scales, we used the sum of ASERT depression items (questions 1 to 4, denoted ASERT_DEP), the sum of ASERT mania items (questions 5 to 8, denoted ASERT_MAN). In line with the original publication, as well as with the results of the internal structure evaluation (below), the non-specific items (questions 9 and 10) were both added to the depression component (denoted ASERT_DEPNSP) or evaluated separately (denoted ASERT_NSP).

The **internal structure** of the English version of the ASERT questionnaire was evaluated using the principal component analysis (PCA) and Cronbach’s alpha^36^, in line with the original publication^20^.

To investigate the relationship between the objective actigraphy-based parameters and the ASERT, as well as the MADRS, YMRS, FAST and BRIAN scales, we extracted the actigraphy parameters from a 14-day time window, ending on the day the questionnaires and scales were assessed. The comparisons were performed using the mixed-effect models with random intercepts for individual participants, using the methodology outlined above.

#### Relapse detection from ASERT and actigraphy data

Next, we investigated the possibility of detecting mood episodes from data obtained remotely. This would offer the possibility to detect clinical changes before they could be detected during regularly scheduled visits. To evaluate the potential utility of weekly ASERT questionnaires and circadian and sleep features extracted from actigraphy for relapse detection, we extracted these features in time windows preceding each clinical rating. A depressive episode (relapse) was defined as MADRS ≥ 15, and a manic relapse as YMRS ≥ 15^37^. As no participants experienced manic episodes, we only evaluated models for depression.

For clinical episode detection, we used the supervised PCA (sPCA) prediction algorithm by Bair et al.^38^. This method was devised to overcome an issue with a high count of possibly correlated features measured on a few individuals. Predictor variables included ASERT responses and actigraphy features, collected up to three weeks prior to the MADRS assessment. The ASERT-based predictors comprised the last ASERT response (within 7 days before the MADRS assessment) and changes in each ASERT item or subscore from the previous ASERT responses.

The actigraphy-based model features were extracted from non-overlapping 7-day windows, and a 14-day window prior to the MADRS assessment. Additionally, differences in actigraphy parameters between the two 7-day windows were added. A comprehensive list of all features selected through the sPCA procedure is provided in Table S5.

The detection model based on the sPCA procedure followed these steps:

1. Preprocessing: Before analysis, each feature was standardized by calculating its Z-score, ensuring uniformity across variables for subsequent analysis.
2. Selection of Relevant Features: Individual features were assessed as predictors of MADRS scores using univariate linear models. Features exhibiting a statistically significant relationship (p-value < 0.05) were retained for further analysis.
3. Summary Model and Components Selection: PCA was conducted on the selected features (see Table S5), generating principal components. Linear models were then fitted using the first 1 to 3 principal components. Components displaying significant association with the outcome were incorporated into the final model.

The model evaluation was performed using the leave-one-subject-out (LOO) approach. The model was iteratively trained (step 3 above) on a dataset comprising all but one patient subsequently generating predicted outcomes for the excluded individual. The training consisted of obtaining new PCA components together with new coefficients for components selected in the Summary model. This process was repeated for each participant, ensuring that each individual served as a test case exactly once.

The procedure was repeated for three different models: i) ASERT-based features only, ii) actigraphy-based features only, and iii) combination of ASERT and actigraphy. The models were evaluated for threshold on predicted MADRS score, defining relapse as MADRS ≥ 15.

## Results

### Overview of study data

A total of 27 individuals with BD were enrolled in the study. Seven patients withdrew their consent due to reluctance to wear the actigraphy wristband for an extended period, particularly during periods of depression, or preference for using a smartwatch as a monitoring device. Data from the remaining 20 participants with follow-up assessments were used for the analysis. The duration of follow-up was between 2 and 11 months (mean 6.6, SD 3.1). An overview of the demographic data of the included participants is provided in Table 1.

**Table 1.** Overview of the study sample.

|  |  |
| --- | --- |
| N | 20 |
| Gender female, n (%) | 15 (75) |
| Age at enrollment, mean (SD) | 36.8 (9.0) |
| Age at first mood symptoms, mean (SD) | 20.5 (7.0) |
| Valid actigraphy data, months, mean (SD) | 6.6 (3.1) |
| Diagnosis, n (%) | BD-I: 7 (35)<br>BD-II: 10 (50)<br>BD type not otherwise specified: 3 (15) |
| MADRS at enrollment, mean SD) | 6.2 (3.5) |
| YMRS at enrollment, mean (SD) | 4.1 (3.2) |
| History of suicide attempts, n (%) | 6 (30) |
| Education, n (%) | Secondary education (High school): 7 (35)<br>Undergraduate education (Bachelor or college completed): 11 (55)<br>Graduate education (Master completed): 2 (10) |
| Medication, n (%) | Mood stabilizer: 13 (65)<br>Antidepressant: 10 (50)<br>Atypical Antipsychotic: 12 (60) |

### Convergent validity of the ASERT relative to clinical scales

The convergent validity of the ASERT relative to the MADRS and YMRS clinical scales was tested using a matched dataset. From the 130 clinician ratings, 116 (from 19 participants) could be matched to ASERT responses within a +/-7 day window. In the matched set, the time differences between ASERT and clinical scale assessments ranged from -4 days (ASERT completed four days before clinical assessment) to +7 days, with a mean of +0.18 days and a SD of 2.27 days.

The results of the LME models with random intercept, comparing clinician ratings with the corresponding subscores of the ASERT, are presented in Table S1. The ASERT depression subscores were significantly associated with MADRS total scores (β=1.42, p<0.001), and the ASERT mania subscores were significantly associated with YMRS total scores (β=0.38, p<0.001). Adding the two non-specific items (9 and 10) to ASERT depression subscores did not significantly improve the model fit (p=0.125, likelihood-ratio test).

### Internal structure of the ASERT

The internal structure of the ASERT questionnaire was validated using the PCA (Table S2). As shown in the table, the questionnaire contains two main components explaining 73% of the overall variance. The PC1, accounting for 45% of the explained variance, is composed of depression and non-specific questions with approximately equal contributions. The PC2, accounting for 28% of the explained variance, is primarily composed of the mania questions, with a small contribution from the non-specific questions.

We evaluated the intended item groups’ reliability using the standardized Cronbach’s alpha coefficient. The reliability was high for the depression items group (Q1:Q4, α=0.914), as well as for the mania items group (Q5:Q8, α=0.865). The summation of depression and non-specific question groups (Q1:Q4, Q9:Q10), which were strongly represented in PC1, also led to a very high reliability of α=0.909.

While the sum of depression and non-specific questions also provides high reliability and convergent validity with the MADRS, we used the more parsimonious variant with four depression questions only due to the more straightforward interpretability and symmetry with the manic part of the ASERT.

### Relationship between ASERT, functional outcome and circadian disruption

For this comparison, we selected the ASERT questionnaire completed closest to the date of the FAST and BRIAN assessments (within a maximum distance of 7 days). The matched set included 112 and 113 ASERT assessments from 19 participants for FAST and BRIAN, respectively. The results of the LME models, comparing the manic and depressive ASERT to both scales, are shown in Supplementary Tables S3a, and S3b. The depression ASERT correlated with FAST total scores (β=1.18, p <0.001) and BRIAN total scores (β=1.20, p<0.001) but not with BRIAN rhythm score (β=0.01, p=0.695). Conversely, the mania ASERT correlated negatively with FAST total scores (β=-0.98, p=0.012), showed no correlation with BRIAN total scores (β=-0.29, p=0.380), but correlated positively with BRIAN rhythm score (β=0.08, p=0.021).

### Actigraphy vs Clinical Scales and ASERT

Actigraphy data collected within a 14-day window before the participants’ assessment by clinical scales was available for 73 pairs of actigraphy features and clinical scales (for MADRS and YMRS) and 72 pairs (for BRIAN and FAST) from 17 participants. The correlation was analyzed using mixed-effects models with random intercepts. While all scales showed the same direction of association with the actigraphy features, except for the BRIAN RHYTHM, the strength and significance of the associations varied, as shown in Figure 2. The MADRS was significantly correlated with all of the investigated actigraphy features except for the acrophase: higher MADRS scores correlated with longer sleep duration, higher IV, lower amplitude and mesor, as well as lower IS. The YMRS was not significantly associated with any of the investigated features, possibly due to the low variability of the YMRS score. BRIAN was most strongly associated with lower IS and lower mesor. The BRIAN RHYTHM component was associated only with a later acrophase. FAST showed no significant association with actigraphy in this comparison.

**Figure 1.**
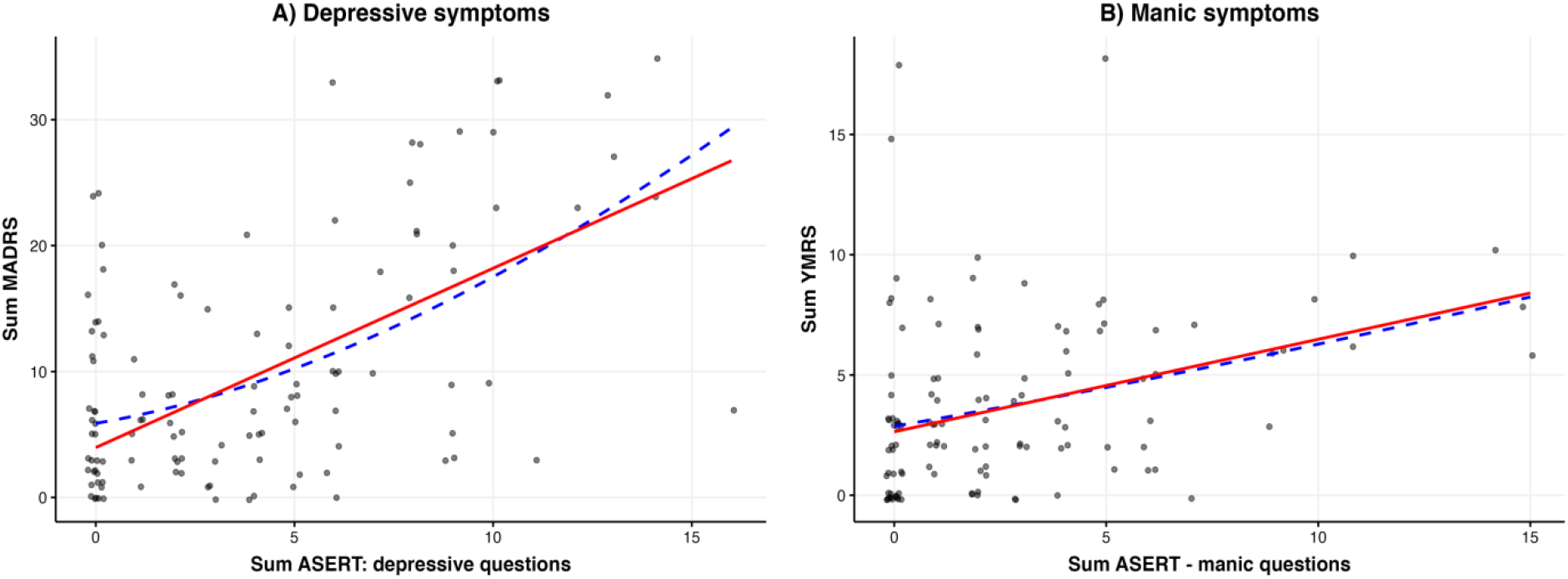
Internal structure of the ASERT. Correlation between the clinical scales and the corresponding ASERT questionnaire subscores. (A) the sum of four questions on depression subscore correlated with the sum of MADRS scores and (B) the sum of four questions on mania subscore correlated with the sum of YMRS scores. For this visualization only, the plotted values include random jitter in both the ASERT questionnaire and MADRS or YMRS. The solid red line represents the fixed-effects part of the relevant mixed-effects model, and the blue dashed line represents a smooth trend (locally estimated scatterplot smoothing). *ASERT: Aktibipo Self-rating; MADRS: Montgomery-Åsberg Depression Rating Scale; YMRS: Young Mania Rating Scale*

**Figure 2.**
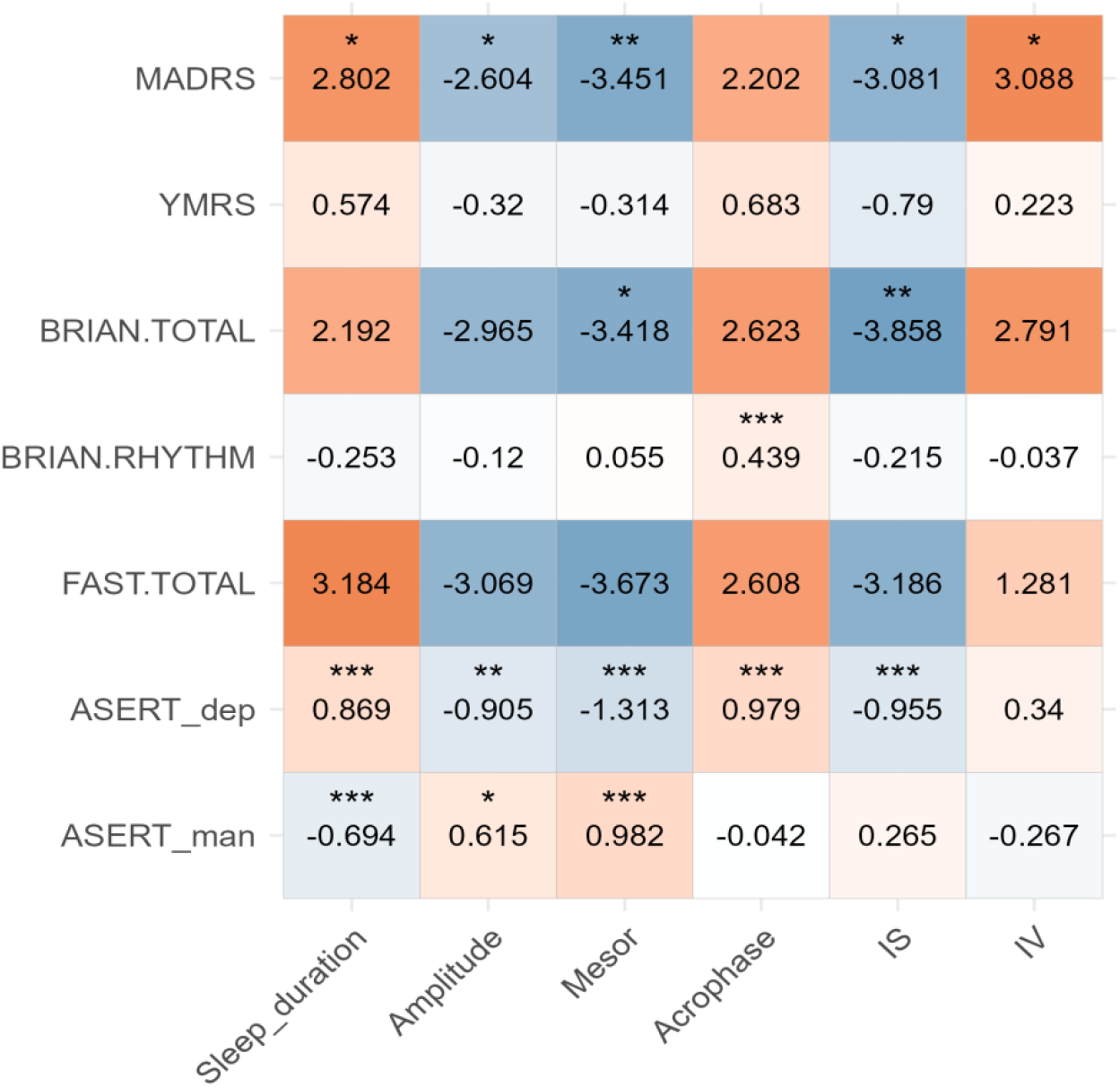
Association between clinical scales, ASERT and actigraphy features in a 14-day window prior to participant’s clinical assessment. The beta coefficients for z-transformed input actigraphy features are shown together with significance levels (* <0.05, ** < 0.01 and *** <0.001).

For the correlation of ASERT subscores with actigraphy features, we used a similar methodology as in the previous analysis^20^, focusing on actigraphy features collected within two weeks before the ASERT self-assessment. This analysis included 343 pairs of completed ASERTs and actigraphy measurements from 18 participants. The depressive ASERT was significantly associated with longer sleep duration, lower amplitude, mesor and IS, and a later acrophase. Higher manic ASERT scores were associated with shorter sleep duration, a higher amplitude and mesor. More detailed results are shown in Supplementary Table S4.

### Relapse detection from ASERT and actigraphy data

For the depression relapse detection, we included all required features, including ASERT responses and complete actigraphy data from one and two weeks before the reference MADRS assessment. This analysis included 65 MADRS assessments from 16 participants with at least one depressive relapse, with an average of four assessments per participant (SD 2.5). Out of these assessments, 50 indicated non-relapse (MADRS < 15) and 15 indicated a depression relapse. Supplementary Table S5 shows relevant features and their PCA component weights for the relevant PCA components. Three models based on different feature sets were evaluated: a) ASERT only, b) actigraphy only, and c) ASERT and actigraphy features combined.

All three models detected depression relapses - see Table 2. The least accurate model uses actigraphy only (62.3% balanced accuracy, 26.7% sensitivity). The model used two PCA components, where the first included mainly physical activity and circadian rhythm-related features, while the second component included sleep regime describing features. The model using solely ASERT-based features achieved higher accuracy (73.7% balanced accuracy, 53.3% sensitivity), again using two PCA components without clear-cut interpretation of their. The combined model, using both ASERT and actigraphy, achieved the best results (81.3% balanced accuracy, 66.7% sensitivity). This model used three PCA components, where the first is mainly based on ASERT scores, the second on circadian rhythm and physical activity, and the third is a combination of ASERT and sleep features (refer to Supplementary Table S5 for details).

**Table 2.** The relapse detection test-set results for ASERT and Actigraphy.

| Model/predictors | Accuracy | Balanced accuracy | Sensitivity | Specificity |
| --- | --- | --- | --- | --- |
| ASERT | 84.6 | 73.7 | 53.3 | 94.0 |
| Actigraphy | 81.5 | 62.3 | 26.7 | 98.0 |
| ASERT & Actigraphy | 89.2 | 81.3 | 66.7 | 96.0 |

## Discussion

In this study, we evaluated the psychometric properties of the English version of the ASERT questionnaire on a North American sample with a comparatively long follow-up of up to 11 months. We replicated the findings of content and convergent validity, the internal structure, and the consistency of the ASERT questionnaire, previously shown using the Czech version and a large European dataset^20^.

Our results confirmed the ASERT’s two-factor structure, aligning with the original study. While the structure of the weaker components (PC3-PC10) differed slightly, this is primarily due to the minimal variance explained by these components, making them more susceptible to noise and random variation. In addition to validating the ASERT’s internal structure, we demonstrated its convergent validity by comparing its depression and mania items to corresponding clinical scales. The strong correlations between MADRS and YMRS with their respective ASERT counterparts support ASERT’s reliability for monitoring mood fluctuations and symptom severity. While the correlation between ASERT mania and YMRS was weaker than in our previous study, this could be attributed to the limited number of manic episodes observed in the current sample.

We found significant associations when correlating the ASERT questionnaire with clinical scales assessing overall functioning (FAST) and biological rhythms (BRIAN). Depression questions on the ASERT (and MADRS) were positively correlated with higher FAST and BRIAN total scores, indicating a link between depression and impairments in overall functioning and biological rhythm. Conversely, mania questions on the ASERT were solely correlated with the BRIAN circadian rhythm component, suggesting a connection between higher mania scores and increased instability in circadian rhythms. We also observed a weak but significant negative association between mania ASERT and FAST, implying that higher self-assessed symptoms of mania might be linked to improved functioning in our sample. This could be due to the limited number of manic episodes in our sample, thus causing mania ASERT, potentially capturing a sense of well-being rather than severe manic symptoms within the subsyndromal states.

BRIAN has proven to be a valuable tool for evaluating biological rhythm dysfunctions associated with mood disorders. Previous research has supported BRIAN’s face validity, showing its correlation with the severity of depressive symptoms, functioning impairment, and poorer quality of life in BD^39–41^. A study by Allega et al.^42^ evaluating objective sleep and activity parameters in BD found that higher BRIAN scores correlated with poor sleep quality indicators, such as wake after sleep onset and total activity count during sleep^42^. Furthermore, individuals with BD showed a higher objective mean nighttime activity, longer total sleep time, and a higher circadian quotient than healthy individuals^43^. These findings align with the BRIAN’s assessment of disrupted biological rhythms in BD.

When comparing data from clinical scales (and subjective assessments) with actigraphy data (objective assessments of activity and sleep), we found a significant association between the stability of daily routines and overall functioning. Patients with lower variability in their daily routines exhibited significantly higher overall functioning. This finding supports the validity of using actigraphy for long-term monitoring of BD. Our previous research has consistently demonstrated that individuals with BD experience sleep disturbances and circadian rhythm dysfunction, including delayed sleep timing, irregular sleep-wake schedules, inconsistent sleep duration, and reduced motor activity^20,26^. These circadian dysfunctions are linked to disrupted daily routines, which can be effectively monitored using actigraphy. Our study further highlights the direct association between these circadian dysfunctions and mood fluctuations and episodes in BD.

Changes in sleep and biological rhythms often precede mood changes in BD, potentially serving as early indicators of relapse. Multiple studies have supported this association. Bauer et al.^44^ found that alterations in sleep-wake patterns often occur before mood shifts during the course of BD. Ng et al.^45^ identified irregular sleep-wake cycles in remitted patients as a risk factor for relapse. Takaesu et al.’s^46^ prospective study further demonstrated that comorbid circadian rhythm sleep-wake disorders in remitted patients can predict shorter time to relapse. In a longitudinal study spanning two years, Gershon et al.^47^ observed that abnormal sleep duration in recovered patients may also serve as a predictor for accelerated depressive recurrences. These findings highlight the importance of addressing sleep disturbances and circadian rhythm dysfunction as important targets for relapse prevention in BD.

Our study highlights the importance of a multi-modal approach to assessing mood relapse, combining subjective self-reports (ASERT) and objective actigraphy data. This approach helps mitigate the potential biases in relying solely on self-reported data. Using a limited dataset, we achieved a sensitivity of 67% (balanced accuracy of 81%) in detecting depression relapse based on a combination of ASERT responses and actigraphy data. Due to the sample size limitations, these results require further validation. All models exhibited high specificity (over 90%), indicating a low false positive rate. However, sensitivity was relatively low. The decision to prioritize sensitivity over specificity is a design choice, and higher sensitivity achieved at the cost of lower specificity can lead to more false alarms. While the ASERT-based features were the main predictor, incorporating actigraphy data improves classification success, increasing balanced accuracy from 74% to 81%. This highlights the complementary nature of these two assessments.

This longitudinal study evaluated multidimensional patient assessments, including self-reports, clinician-administered scales, and objective actigraphy monitoring. This approach provides a rich description of patients’ mood status and disease course. Each assessment offers a unique perspective on a patient’s current symptoms.While subjective and potentially influenced by a patient’s insight during symptomatic periods, self-reports can contribute positively to adherence when questionnaires are easily completed, such as the ASERT via a mobile app. This allows for frequent, long-term symptom monitoring at a low cost and without requiring clinician involvement. This study validated the ASERT questionnaire in a distinct population, demonstrating its utility in accurately assessing mood fluctuations in BD, even with a smaller sample size. The ASERT emerges as a valuable tool for remote mood monitoring in research and clinical settings.

Interpreting our findings, we need to acknowledge their limitations. While the sample size is comparable to the previous actigraphy studies^10^, it still may have limited the statistical power. Further, all participants were taking prescribed medications, as treatment discontinuation would not be ethical. Yet, psychotropic medications are known to influence sleep and circadian rhythms^48,49^. Also, the prevalence of comorbidities in BD is high^50^ which could have influenced circadian rhythms. Finally, the relatively small number of depressive and manic episodes observed during the follow-up period may have constrained the accuracy of relapse detection. Despite the study’s limitations, its longitudinal design with a follow-up up to 11 months and frequent weekly assessments helps better understand the relationship between sleep, circadian rhythms, and mood changes in BD.

Given the recurrent nature of BD and its susceptibility to routine triggers, long-term monitoring is crucial for maintaining stable clinical outcomes and functioning. This study demonstrates that long-term stability of sleep and activity assessed by actigraphy reflects better clinical outcomes and healthier overall functioning. By utilizing digital tools, clinicians can remotely assess patients, gain a deeper understanding of their illness course, and make more informed treatment decisions. This approach holds the potential to reduce adverse outcomes, such as mood episode recurrence and suicide risk, thus enhancing the overall quality of life for individuals with BD. A key area for future research is the development of accurate tools to monitor the course of illness and inform effective disease management. Data-driven analyses incorporating clinically relevant metrics like sleep, physical activity, circadian rhythmicity, and mood assessments can support targeted interventions. This research has the potential to significantly reduce the burden of BD on individuals, families, and society.

## Data Availability

Data produced in the present study are available upon reasonable request to the authors.

## Acknowledgements

Work on this study was partially supported by the Mitacs Accelerate International Award (#IT23407), by the Ministry of Health of the Czech Republic, grant nr. NU23-04-00534 and the ERDF-Project Brain dynamics, No. CZ.02.01.01/00/22_008/0004643. The Kapczinski lab was supported by the Strategic Alignment Fund/McMaster University and the St. Joseph’s Healthcare Foundation, Hamilton, Canada, and FAPERGS (21/2551-0001990-5), RENASAM (445154/2023-3) and INCT-TM (465458/2014-9), Porto Alegre, Brazil.

