## Supplementary information for "Self-assessment and rest-activity rhythm monitoring for effective bipolar disorder management: a longitudinal actigraphy study"

To the paper:

### 1. Convergent validity: ASERT vs YMRS and MADRS clinical scales

Table S1 shows linear mixed-effect random intercept model results, comparing the ASERT subscores to the respective clinical scale summary scores. ASERT responses were obtained within 7 days from the clinical scale. Significance testing was done using the Satterwaite's method (lmerTest R package).

**Table S1.** Comparison of ASERT subscores to relevant clinical scales

| | **YMRS LME vs ASERT_MAN** | | | **MADRS LME vs ASERT_DEP** | | | **MADRS LME vs ASERT_DEPNSP** | | |
|---|---|---|---|---|---|---|---|---|---|
| *Predictors* | *Estimates* | *CI* | *p* | *Estimates* | *CI* | *p* | *Estimates* | *CI* | *p* |
| (Intercept) | 2.64 | 1.66 – 3.62 | <0.001 | 3.96 | 1.38 – 6.54 | 0.003 | 2.75 | 0.01 – 5.49 | 0.049 |
| **ASERT mania** | **0.38** | **0.18 – 0.59** | **<0.001** | | | | | | |
| **ASERT depression** | | | | **1.42** | **1.05 – 1.80** | **<0.001** | | | |
| **ASERT depression + non-specific** | | | | | | | **1.04** | **0.77 – 1.31** | **<0.001** |
| **Random Effects** | | | | | | | | | |
| $\sigma^2$ | 10.23 | | | 48.54 | | | 47.74 | | |
| $\tau_{00}$ | 1.11 ID | | | 10.94 ID | | | 10.73 ID | | |
| ICC | 0.10 | | | 0.18 | | | 0.18 | | |
| N | 19 ID | | | 19 ID | | | 19 ID | | |
| Observations | 116 | | | 116 | | | 116 | | |
| Marginal $R^2$ / Conditional $R^2$ | 0.124 / 0.210 | | | 0.350 / 0.469 | | | 0.371 / 0.487 | | |

### 2. Internal structure of the ASERT

Table S2 shows the internal structure of the ASERT responses, as analyzed on all 531 ASERT responses.

**Table S2**. Internal structure of the ASERT questionnaire resulting from the PCA analysis

| | **#** | **Item text** | **PC1** | **PC2** | **PC3** | **PC4** | **PC5** | **PC6** | **PC7** | **PC8** | **PC9** | **PC10** |
|---|---|---|---|---|---|---|---|---|---|---|---|---|
| **depression** | 1 | I feel sad, downhearted | 0.418 | -0.130 | 0.196 | -0.311 | 0.108 | -0.286 | 0.079 | -0.612 | -0.284 | -0.345 |
| | 2 | I do not enjoy anything, and nothing pleases me | 0.371 | -0.092 | 0.275 | 0.059 | -0.043 | -0.072 | -0.080 | -0.203 | 0.781 | 0.332 |
| | 3 | I have no energy | 0.396 | -0.172 | -0.040 | 0.477 | 0.278 | -0.181 | -0.576 | 0.173 | -0.305 | 0.140 |
| | 4 | I feel gloomy and pessimistic about the future | 0.415 | -0.083 | 0.438 | -0.129 | -0.406 | -0.036 | 0.267 | 0.579 | -0.200 | 0.002 |
| **mania** | 5 | I feel unusually great, optimistic | -0.058 | 0.449 | -0.012 | 0.270 | -0.433 | -0.449 | -0.231 | 0.018 | 0.189 | -0.493 |
| | 6 | I have excess energy | -0.026 | 0.506 | 0.098 | 0.094 | -0.202 | -0.184 | 0.117 | -0.290 | -0.342 | 0.660 |
| | 7 | My thinking is very fast, others cannot keep up with me | 0.158 | 0.496 | 0.045 | -0.039 | 0.688 | -0.213 | 0.318 | 0.285 | 0.133 | -0.087 |
| | 8 | I need to sleep less than usual | 0.061 | 0.430 | 0.399 | -0.200 | 0.056 | 0.606 | -0.467 | -0.054 | -0.048 | -0.138 |
| **non-specific** | 9 | I feel restless, tense | 0.370 | 0.175 | -0.646 | -0.545 | -0.156 | -0.030 | -0.228 | 0.141 | 0.059 | 0.131 |
| | 10 | I cannot focus | 0.436 | 0.140 | -0.323 | 0.484 | -0.125 | 0.480 | 0.382 | -0.178 | -0.012 | -0.168 |
| | | **Standard deviation** | 2.352 | 1.847 | 0.880 | 0.773 | 0.707 | 0.640 | 0.612 | 0.517 | 0.453 | 0.423 |
| | | **Proportion of Variance** | 0.451 | 0.279 | 0.063 | 0.049 | 0.041 | 0.033 | 0.031 | 0.022 | 0.017 | 0.015 |
| | | **Cumulative Proportion** | 0.451 | 0.730 | 0.793 | 0.842 | 0.883 | 0.916 | 0.947 | 0.969 | 0.985 | 1.000 |

### 3. ASERT vs BRIAN and FAST

Tables S3a and S3b provide an overview of linear mixed effect model parameters for comparison of the BRIAN and FAST scales (dependent variable) predicted by responses to the *depressive* and *manic* part of the ASERT, respectively.

**Table S3a.** ASERT_DEP vs FAST and BRIAN

| | FAST.TOTAL | | | BRIAN.TOTAL | | | BRIAN.RHYTHM | | |
|---|---|---|---|---|---|---|---|---|---|
| *Predictors* | *Estimates* | *CI* | *p* | *Estimates* | *CI* | *p* | *Estimates* | *CI* | *p* |
| (Intercept) | 14.33 | 8.48 – 20.18 | **<0.001** | 31.36 | 27.89 – 34.83 | **<0.001** | 6.31 | 5.74 – 6.88 | **<0.001** |
| sum quest dep | 1.18 | 0.72 – 1.63 | **<0.001** | 1.20 | 0.81 – 1.58 | **<0.001** | 0.01 | -0.04 – 0.06 | 0.695 |
| **Random Effects** | | | | | | | | | |
| $\sigma^2$ | 55.07 | | | 43.93 | | | 0.63 | | |
| $\tau_{00}$ | 134.93 $_{ID}$ | | | 35.68 $_{ID}$ | | | 1.22 $_{ID}$ | | |
| ICC | 0.71 | | | 0.45 | | | 0.66 | | |
| N | 19 $_{ID}$ | | | 19 $_{ID}$ | | | 19 $_{ID}$ | | |
| Observations | 112 | | | 113 | | | 113 | | |
| Marginal $R^2$ / Conditional $R^2$ | 0.102 / 0.740 | | | 0.224 / 0.572 | | | 0.001 / 0.658 | | |

**Table S3b.** ASERT_MAN vs FAST and BRIAN

| | FAST.TOTAL | | | BRIAN.TOTAL | | | BRIAN.RHYTHM | | |
|---|---|---|---|---|---|---|---|---|---|
| *Predictors* | *Estimates* | *CI* | *p* | *Estimates* | *CI* | *p* | *Estimates* | *CI* | *p* |
| (Intercept) | 22.43 | 15.93 – 28.93 | **<0.001** | 37.42 | 33.48 – 41.37 | **<0.001** | 6.09 | 5.58 – 6.60 | **<0.001** |
| sum quest man | -0.98 | -1.74 – -0.22 | **0.012** | -0.29 | -0.93 – 0.36 | 0.380 | 0.08 | 0.01 – 0.16 | **0.021** |
| **Random Effects** | | | | | | | | | |
| $\sigma^2$ | 63.88 | | | 58.63 | | | 0.64 | | |
| $\tau_{00}$ | 163.01 $_{ID}$ | | | 44.66 $_{ID}$ | | | 0.87 $_{ID}$ | | |
| ICC | 0.72 | | | 0.43 | | | 0.58 | | |
| N | 19 $_{ID}$ | | | 19 $_{ID}$ | | | 19 $_{ID}$ | | |
| Observations | 112 | | | 113 | | | 113 | | |
| Marginal $R^2$ / Conditional $R^2$ | 0.044 / 0.731 | | | 0.009 / 0.437 | | | 0.049 / 0.597 | | |

### 4. ASERT and clinical scales vs actigraphy

Table S4 summarizes the mixed-effect model results for comparison between actigraphy, ASERT and clinical scales. Beta slope coefficients(β1) are calculated for scaled (z-transformed) predictor data for comparability. The $\varepsilon^2$ are the estimated epsilon squared effect size values.

**Table S4.** Results of LME models comparing actigraphy to clinical scales and ASERT

| **predictor** | **Sleep_duration** | | | **Amplitude** | | | **Mesor** | | | **Acrophase** | | | **IS** | | | **IV** | | |
|---|---|---|---|---|---|---|---|---|---|---|---|---|---|---|---|---|---|---|
| **dep. variable** | **β1** | **P** | **ε2** | **β1** | **P** | **ε2** | **β1** | **P** | **ε2** | **β1** | **P** | **ε2** | **β1** | **P** | **ε2** | **β1** | **P** | **ε2** |
| MADRS | **2.802** | **0.023** | 0.107 | **-2.604** | **0.039** | 0.132 | **-3.451** | **0.006** | 0.259 | 2.202 | 0.073 | 0.061 | **-3.081** | **0.012** | 0.153 | **3.088** | **0.012** | 0.289 |
| YMRS | 0.574 | 0.160 | 0.014 | -0.320 | 0.436 | 0.000 | -0.314 | 0.445 | 0.000 | 0.683 | 0.116 | 0.058 | -0.790 | 0.052 | 0.038 | 0.223 | 0.587 | 0.000 |
| BRIAN.TOTAL | 2.192 | 0.120 | 0.029 | -2.965 | 0.053 | 0.076 | **-3.418** | **0.032** | 0.112 | 2.623 | 0.057 | 0.051 | **-3.858** | **0.005** | 0.122 | 2.791 | 0.068 | 0.062 |
| BRIAN.RHYTHM | -0.253 | 0.066 | 0.034 | -0.120 | 0.442 | 0.000 | 0.055 | 0.735 | 0.000 | **0.439** | **0.001** | 0.167 | -0.215 | 0.109 | 0.023 | -0.037 | 0.812 | 0.000 |
| FAST.TOTAL | 3.184 | 0.058 | 0.036 | -3.069 | 0.104 | 0.027 | -3.673 | 0.059 | 0.045 | 2.608 | 0.121 | 0.020 | -3.186 | 0.050 | 0.039 | 1.281 | 0.498 | 0.000 |
| ASERT_dep | **0.869** | **0.000** | 0.035 | **-0.905** | **0.002** | 0.029 | **-1.313** | **0.000** | 0.059 | **0.979** | **0.000** | 0.077 | **-0.955** | **0.000** | 0.046 | 0.340 | 0.205 | 0.002 |
| ASERT_man | **-0.694** | **0.001** | 0.031 | **0.615** | **0.011** | 0.016 | **0.982** | **0.000** | 0.044 | -0.042 | 0.858 | 0.000 | 0.265 | 0.178 | 0.002 | -0.267 | 0.235 | 0.001 |

### 5. ASERT and actigraphy-based predictors of depressive relapse

Table S5 summarizes the ASERT-based and actigraphy-based features, used for depression prediction and resulting principal components of the supervised PCA procedure that were used in the model. Three different models and thus three different PCA components are compared. The features marked _*diff* correspond to differences between feature values from subsequent weeks.

**Table S5.** Predictive features for depressive relapse prediction

| | ASERT only | | Actigraphy only | | Combined | | |
|---|---|---|---|---|---|---|---|
| **Relevant features** | **PC1** | **PC2** | **PC1** | **PC2** | **PC1** | **PC2** | **PC3** |
| asert_q_1 | 0.192 | -0.204 | | | 0.187 | -0.078 | -0.175 |
| asert_q_2 | 0.214 | -0.241 | | | 0.216 | 0.030 | -0.204 |
| asert_q_3 | 0.206 | -0.221 | | | 0.199 | -0.101 | -0.179 |
| asert_q_4 | 0.223 | -0.190 | | | 0.224 | 0.024 | -0.160 |
| asert_q_9 | 0.153 | -0.204 | | | 0.152 | -0.035 | -0.192 |
| asert_q_10 | 0.206 | -0.231 | | | 0.202 | -0.057 | -0.210 |
| asert_fill_time | 0.110 | 0.039 | | | 0.102 | -0.057 | 0.067 |
| asert_sum_dep | 0.241 | -0.246 | | | 0.238 | -0.038 | -0.206 |
| asert_sum_all | 0.244 | -0.255 | | | 0.246 | -0.003 | -0.246 |
| asert_sum_ns | 0.197 | -0.239 | | | 0.195 | -0.051 | -0.220 |
| asert_sum_dep_ns | 0.247 | -0.267 | | | 0.244 | -0.047 | -0.232 |
| asert_q_1_diff | 0.230 | 0.294 | | | 0.223 | -0.020 | 0.287 |
| asert_q_2_diff | 0.285 | 0.316 | | | 0.278 | 0.0268 | 0.323 |
| asert_q_3_diff | 0.246 | 0.187 | | | 0.234 | -0.088 | 0.197 |
| asert_q_4_diff | 0.271 | 0.226 | | | 0.264 | -0.022 | 0.236 |
| asert_q_10_diff | 0.230 | 0.164 | | | 0.225 | -0.036 | 0.148 |
| asert_sum_dep_diff | 0.283 | 0.278 | | | 0.274 | -0.032 | 0.284 |
| asert_sum_ns_diff | 0.224 | 0.158 | | | 0.223 | 0.014 | 0.134 |
| asert_sum_dep_ns_diff | 0.282 | 0.252 | | | 0.275 | -0.017 | 0.246 |
| Mesor_14 | | | 0.355 | -0.317 | 0.073 | 0.339 | -0.059 |
| IS_14 | | | 0.353 | 0.372 | 0.002 | 0.359 | 0.076 |
| IV_14 | | | -0.396 | 0.148 | -0.067 | -0.379 | 0.098 |
| Ampl_14 | | | 0.402 | -0.075 | 0.099 | 0.383 | 0.002 |
| SleepDur_MAD_14 | | | -0.151 | -0.327 | 0.030 | -0.166 | -0.154 |
| Mesor_7 | | | 0.354 | -0.351 | 0.077 | 0.338 | -0.088 |
| IS_7 | | | 0.335 | 0.437 | -0.001 | 0.339 | 0.060 |
| IV_7 | | | -0.396 | 0.159 | -0.057 | -0.379 | 0.129 |
| SleepDur_MAD_7 | | | -0.106 | -0.371 | 0.060 | -0.127 | -0.143 |
| SleepDur_MAD_7_diff | | | 0.056 | 0.388 | -0.068 | 0.078 | 0.162 |
